# COVID-19 vaccine-associated cerebral venous thrombosis in Germany

**DOI:** 10.1101/2021.04.30.21256383

**Authors:** Jörg B. Schulz, Peter Berlit, Hans-Christoph Diener, Christian Gerloff, Andreas Greinacher, Christine Klein, Gabor C. Petzold, Marco Piccininni, Sven Poli, Rainer Röhrig, Helmuth Steinmetz, Thomas Thiele, Tobias Kurth, the DGN SARS-CoV-2 Vaccination Study Group

## Abstract

**Objective:** Reports of cerebral sinus and venous thrombosis (CVT) after ChAdOx1 vaccination against SARS-CoV-2 have raised safety concerns. We aimed to estimate the incidence of CVT within one month from first dose administration and the frequency of vaccine-induced immune thrombotic thrombocytopenia (VITT) as the underlying mechanism after vaccination with BNT162b2, ChAdOx1, and mRNA-1273, in Germany.

**Methods:** A web-based questionnaire was e-mailed to all Departments of Neurology. We asked to report cases of CVT within one month of a COVID-19 vaccination. Other cerebral events could also be reported. Incidence rates of CVT were calculated by using official statistics of nine German States.

**Results:** A total of 45 CVT cases were reported. In addition, 9 primary ischemic strokes, 4 primary intracerebral hemorrhages, and 4 other neurological events were recorded. Of the CVT patients, 35 (77.8%) were female, and 36 (80.0%) were below the age of 60 years. Fifty-three events were observed after vaccination with ChAdOx1 (85.5%), 9 after BNT162b2 (14.5%), and none after mRNA-1273 vaccination. After 7,126,434 first vaccine doses, the incidence rate of CVT within one month from first dose administration was 6.5 (95% CI, 4.4-9.2) per 100,000 person-years for all vaccines and 17.9 (11.8-26.1) for ChAdOx1 (after 2,320,535 ChAdOx1 first doses). The adjusted incidence rate ratio was 9.68 (3.46-34.98) for ChAdOx1 compared to mRNA-based vaccines and 3.14 (1.22-10.65) for women compared to non-women. In 26/45 patients with CVT (57.8%), VITT was graded highly probable.

**Conclusions:** Given an incidence of 0.22–1.75 per 100,000 person-years for CVT in the general population, these findings point towards a higher risk for CVT after ChAdOx1 vaccination, especially for women.

## Introduction

A major breakthrough in managing the COVID-19 pandemic was the development and administration of vaccines against SARS-CoV-2, namely BNT162b2 (BioNTech/Pfizer), mRNA-1273 (Moderna), Ad26.COV2.S (Johnson & Johnson) and ChadOx1 (AstraZeneca). Typical side effects of these vaccines were reported in clinical trials with several thousands of volunteers but without evidence of a vaccine-associated increase in thromboembolic events^1–5^. Until April 2021, several vaccines have been approved and administered to millions of people. In Germany, 16,428,425 persons received the first and 5,517,282 the second dose of a vaccine as of April 18, 2021^6^. These included about 16.2 million BNT162b2 doses, 1.2 million mRNA-1273 doses, and 4.6 million ChAdOx1m doses.

Outside of the context of COVID-19 vaccination, cerebral venous thrombosis is a very rare disease with an incidence of about 0.22 – 1.75 per 100.00 person-years, based on data from four European countries, Australia, Iran, and Hong Kong^7–9^. Well-known risk factors are female sex, pregnancy, infections, and hypercoagulability^10^. Within hypercoagulability, hormone-related and genetic prothrombotic disorders are the most frequent causes^11^. Until the end of March 2021, the majority of persons vaccinated with ChAdOx1 in Germany were below the age of 60 years ^6^. ChAdOx1 was initially only recommended in Germany for persons below the age of 65 due to insufficient data on efficacy and safety among the elderly. In several European countries, cases of cerebral venous thrombosis were reported in temporal relationship with ChAdOx1 vaccine administration. An immune-mediated mechanism termed vaccine-induced thrombocytopenic thrombosis (VITT) has been suggested to underlie these serious adverse events^12–14^. At the beginning of March 2021, 30 venous thromboembolic events were reported to EMA out of about 5 million persons who had received the ChAdOx1 vaccine^15^. At that time, the Danish National Patient Registry did not report a higher incidence of thromboembolic events in the Danish population but excluded cases of sinus-venous thrombosis from their analysis because of low incidence^16^.

The aim of this report is to describe reported cases of cerebrovascular events in temporal relation to COVID-19 vaccination in Germany until April 14, 2021, based on a retrospective survey. We further aim at providing an incidence estimate of cerebral venous thrombosis within 31 days from first vaccine dose administration by vaccine type, age, and sex for nine German states

## Methods

### Data collection

We designed a web-based questionnaire which was e-mailed to all departments of neurology of university (n=40) and non-university (n=251) hospitals in Germany on April 6, 2021. Data collection was closed on April 14, 2021 (24:00). The survey focused on the report of cerebral sinus-venous thrombosis and cerebral venous thrombosis events that had occurred within 31 days after COVID-19 vaccination in 2021. However, the questionnaire also allowed the reporting of other cerebrovascular events in possible temporal relationship with a COVID-19 vaccination. We combined cerebral sinus-venous thrombosis and cerebral venous thrombosis without the involvement of the vena cerebri magna—hereafter referred to as cerebral sinus and/or venous thrombosis (CVT). Thirty-seven (92%) neurology departments at university hospitals (tertiary centers) and 75 (30%) neurology departments of non-university hospitals responded (Figure 1). We recorded information about the type of vaccination, symptoms, coagulation parameters, clinical course, and clinical outcomes. We developed a written protocol for data collection (see appendix). The protocol was approved by the Ethics Committee (Vote-No. 142/21, Ethics Committee of the Medical Faculty at RWTH University). Data protection and privacy conformity have been confirmed by the Data Protection Officer and the Information Security Officer of RWTH Aachen University Hospital. Coagulation parameters were also collected from the respective local laboratories. For a subgroup of patients, anti-Heparin/Platelet Factor 4 Antibody (PF4)/polyanion-IgG EIA and a platelet activation assay were performed in the laboratory of the Institute for Immunology and Transfusion Medicine at the University of Greifswald as described^13^. For the PF4 antibody results, we used information from the central laboratory in Greifswald, and only if missing, we considered test results if positive from the respective local hospitals.

**Figure 1.**
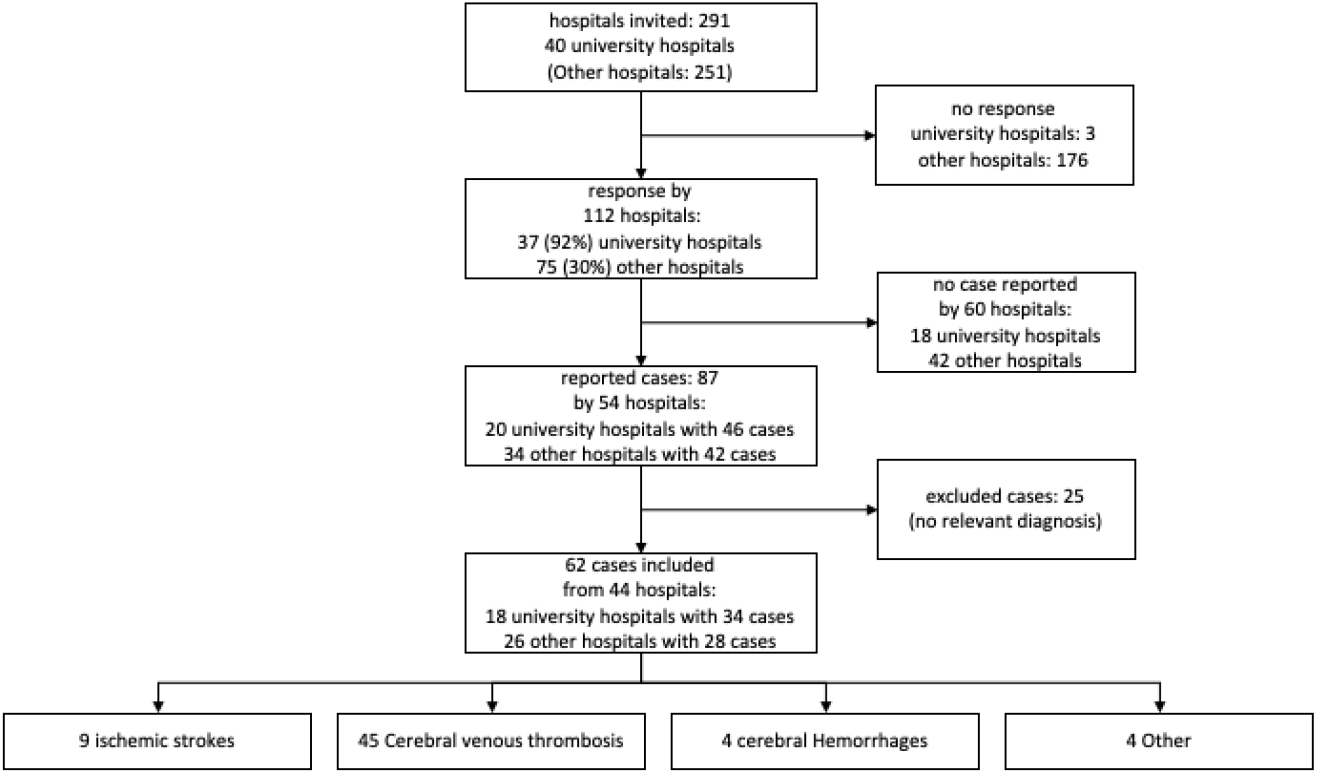
Study Flow Chart.

Based on the first reported cases^12,13^ we devised a grading system using the following criteria in order to classify each event according to its likelihood of being associated to COVID-19 vaccination: (a) time from last vaccine shot administration between one and 16 days, (b) thrombocytopenia (<150/nL) or relative thrombocytopenia (drop of thrombocytes of at least 50%), (c) positive enzyme-linked immunosorbent assay (ELISA) to detect platelet factor 4 (PF4)-polyanion antibodies, (d) positive modified (PF4-enhanced) platelet-activation assay (VITT function test)^13^. Each criterion loaded the score with 1 point. All cases were evaluated in depth by four members of the Task Force. Cases that fulfilled criteria a and b, but no test results were available for c and d, were rated with a score of 2+ to contrast them to those cases with negative results for c and d. A score of 2+ and higher was considered a high grade (highly probable VITT).

### Research Hypotheses

We formulated the following a priori research hypotheses:

1. Vaccine-induced CVTs are restricted to COVID-19 vaccination with ChAdOx1 and do not occur after vaccination with mRNA-based vaccines.
2. Females, particularly below the age of 60 years, are more likely to be diagnosed with CVT after COVID-19 vaccination.
3. Patients with vaccine-induced CVT after COVID-19 vaccination have a high prevalence of antibodies against thrombocytes and/or thrombocytopenia, resulting in venous thrombosis and bleedings.
4. VITT-mediated neurological events are not restricted to vaccine-induced CVTs but may also result in cerebrovascular arterial thrombotic events.

### Statistical analysis

Characteristics of the reported cerebrovascular cases were summarized as frequency and percentage or mean, standard deviation, median, and range for qualitative and quantitative variables, respectively. Descriptive statistics were reported for the overall cases and by subgroups.

In order to compute the incidence rate of CVT within one month from first vaccine shot administration, we divided the number of cases that occurred within 31 days from first vaccine shot administration by the overall amount of person-time spent at risk during the time window of interest.

We obtained CC-BY licensed data from the Robert Koch-Institute (the German National Institute of Public Health) about the number of vaccine shots administered by calendar week, age group, vaccine type, and state separately for only females and for everyone (numbers for non-females were obtained by difference). The number of vaccine shots administered within these subgroups was only available for nine German states, and no distinction was possible between first and second doses.

Therefore, we restricted our estimation of the incidence to the area of the nine German States (Baden-Wuerttemberg, Bremen, Hamburg, Mecklenburg-Western Pomerania, Lower Saxony, North Rhine-Westphalia, Rhineland-Palatinate, Saarland, and Schleswig-Holstein). We assumed that a case originated in this area if the hospital recording it was located in one of the nine States. We only considered cases occurring within 31 days from the first vaccine dose administration. For cases occurring after the second shot, we computed the time from first dose assuming that the second dose was administered 10 weeks, 21 days, 14 days after the first for ChAdOx1, BNT162b2, and mRNA-1273, respectively.

Within every stratum of state, age group (<60, 60+), sex (female, non-female), and vaccine type (ChAdOx1, BNT162b2, and mRNA-1273), we approximated the number of first and second doses administered every calendar week. We assumed individuals receiving their second dose in a given week were the same who had received their first dose a fixed amount of weeks before (10 for ChAdOx1, 3 for BNT162b2, and 2 for mRNA-1273). If the number of attributed second doses in a week was higher than the total registered number of administered doses, the remaining doses were attributed to the following week (and so on, iteratively). The number of first doses was obtained by the difference between the total number of doses and the estimated number of second doses administered in the week.

The number of person-years each vaccinated individual spent at risk during the time window of interest (one month from first vaccine shot administration) was computed as the number of days between the day of the first dose administration (assumed in the middle of Wednesday) and the 31st day after the first dose administration or the end of the study period (April 14, 2021) whichever occurred first, divided by 365.25. We only considered the time contributed by individuals who received their first dose between December 28, 2020, and April 11, 2021.

Overall and group-specific incidence rates were expressed as number of cases per 100,000 person-years and reported along with their 95% exact Poisson confidence intervals.

Our approach relies on the assumptions that no individual moved from a state-age-sex-vaccine group to another during the 31 days following first dose administration, that no competing events occurred during this time window, and that everyone received a second dose of the vaccine according to the above-specified schedule.

Finally, we fitted a Poisson log-linear regression model with the logarithm of the person-years as offset to investigate the association between age group (<60, 60+), sex (female, non-female), vaccine class (ChAdOx1, mRNA-based vaccines), and the CVT incidence rate within one month from first dose administration. No interaction terms were included. P-values lower than or equal to 0.05 were considered statistically significant.

All analyses were performed using R version 4.0.3 and RStudio 1.1.456.

## Results

A total of 291 departments of neurology were contacted, of which 112 reported back (Figure 1). After excluding duplicates, and cases without cerebrovascular outcomes, 62 patients with a cerebral event were reported in close temporal proximity to the vaccination against COVID-19 (Figure 1), of which 45 were CVTs. Reported cases had a mean age of 46.7 years, and 75.8% were female. 6/52 (11.5%) cases were smokers, 3/59 cases (5.1%) were obese, and 1/59 (1.7%) reported a previous thrombosis event (Table 1).

**Table 1.**
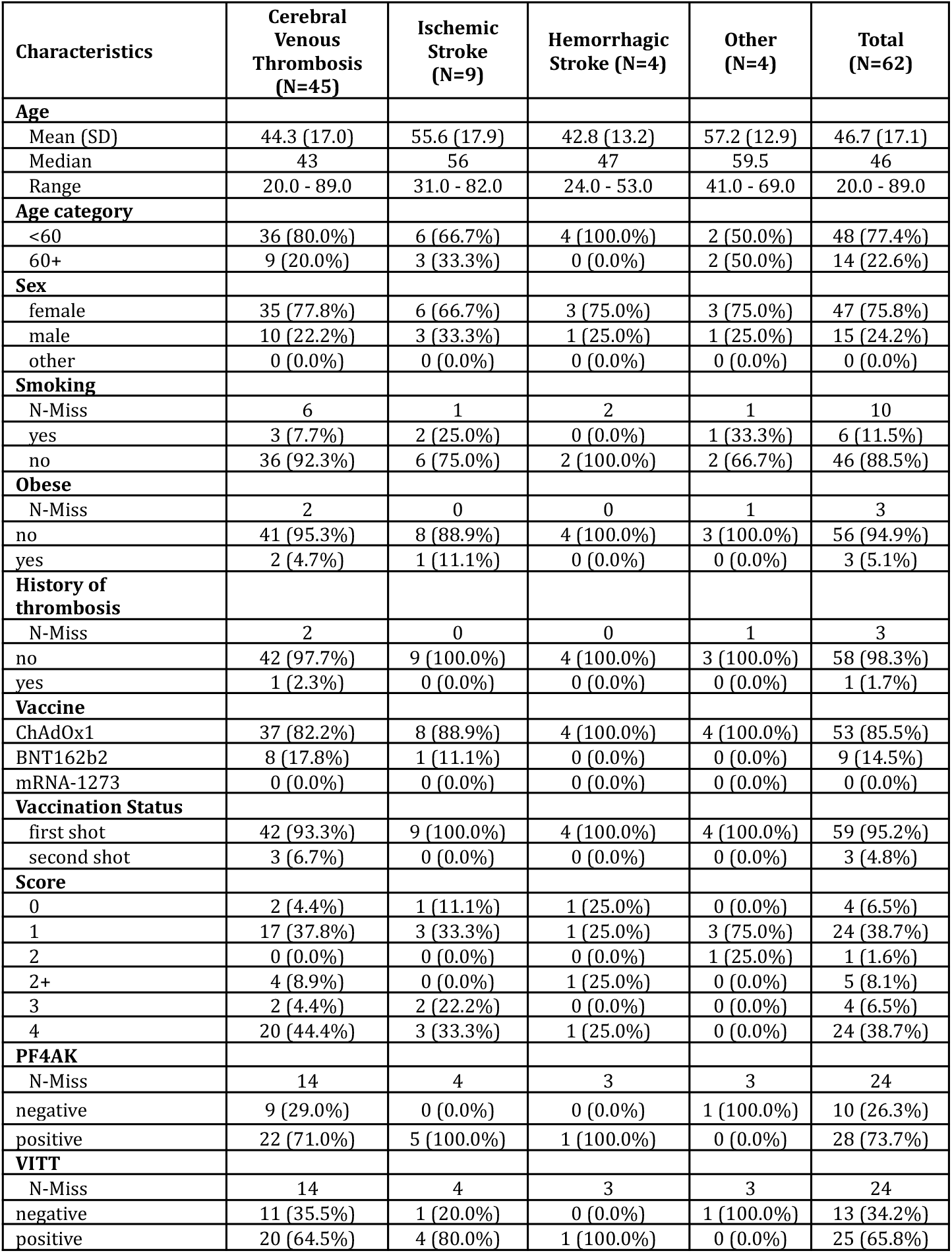
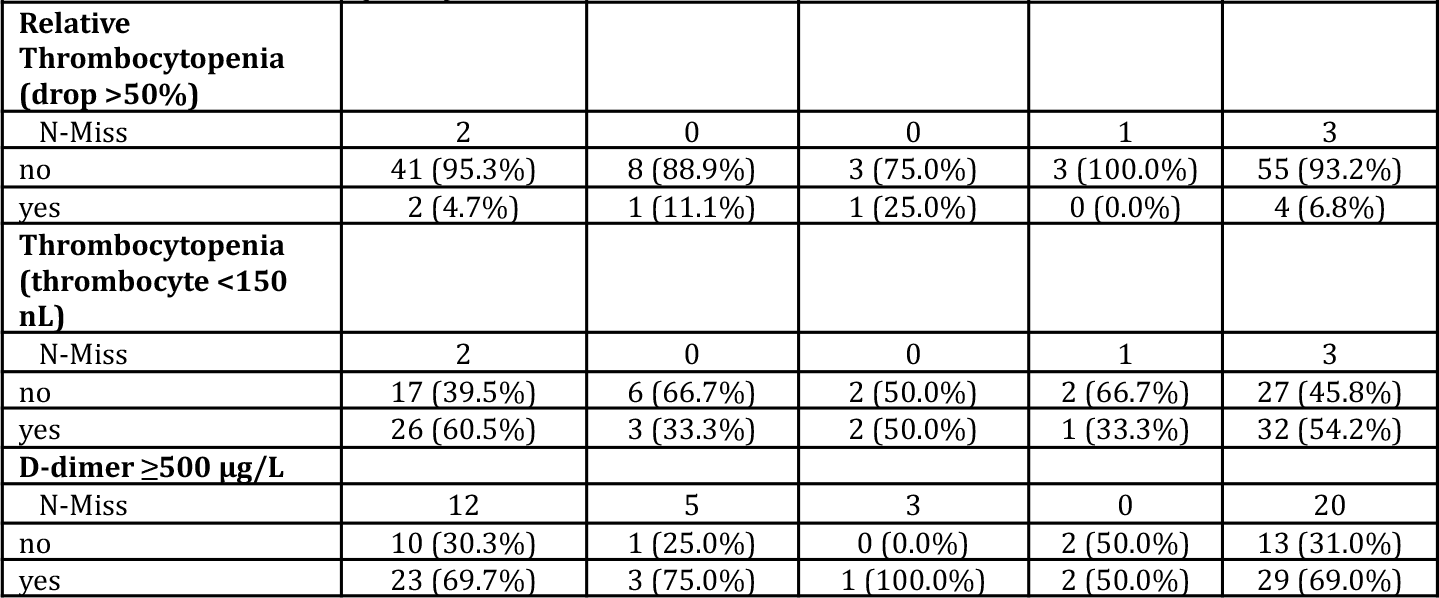
Characteristics of included cases with cerebral and central nervous system events within 31 days from Covid-19 vaccination.

All reported cases occurred after vaccination with ChAdOx1 (85.5%) and BNT162b2 (14.5%). No cases were reported with mRNA-1273. No other vaccines were used in Germany during the study period. The initial diagnosis of CVT was confirmed by MR and MR-venography or CT and CT-venography in all cases. We identified 37 CVT cases after ChAdOx1 and 8 after BNT162b2. Of the 45 patients with CVT, 35 (77.8%) were female, and 36 (80.0%) were below the age of 60 years (Table 1). Primary intracerebral hemorrhages (ICH) were observed in 4 cases, and 9 patients had primary cerebral ischemia (Table 1). In addition, a total of 4 patients were reported with other diagnoses (1 transient global amnesia, 1 spinal artery ischemia, and 2 nausea, one of them with headache).

Two out of 62 patients (3.2%) presented with dermal petechia, two (3.2%) with subdermal hematoma, and two (3.2%) with bleedings in other territories. A total of 59 (95.2%) events occurred after the first dose administration of a vaccine and 3 (4.8%) after the second dose administration (all three BNT162b2) (Table 1). Forty-two (93.3%) and 3 (6.7%) CVT events occurred after first and second dose administration, respectively (Table 1).

Sixty-one cases (98.4%) experienced first neurological symptoms within 31 days (approximated value for cases occurred after BNT162b2 second shot) from the first vaccine shot administration. All events after ChAdOx1 occurred after the first dose, but very few people in Germany received a second dose during the study period. Days from first ChAdOx1 shot to neurological symptoms’ onset are presented in Figure 2. The median time interval from last administered vaccine shot to neurological symptoms was 9 days (range 1 to 25) for CVT events. None of the cases had a previously confirmed SARS-CoV-2-infection.

**Figure 2.**
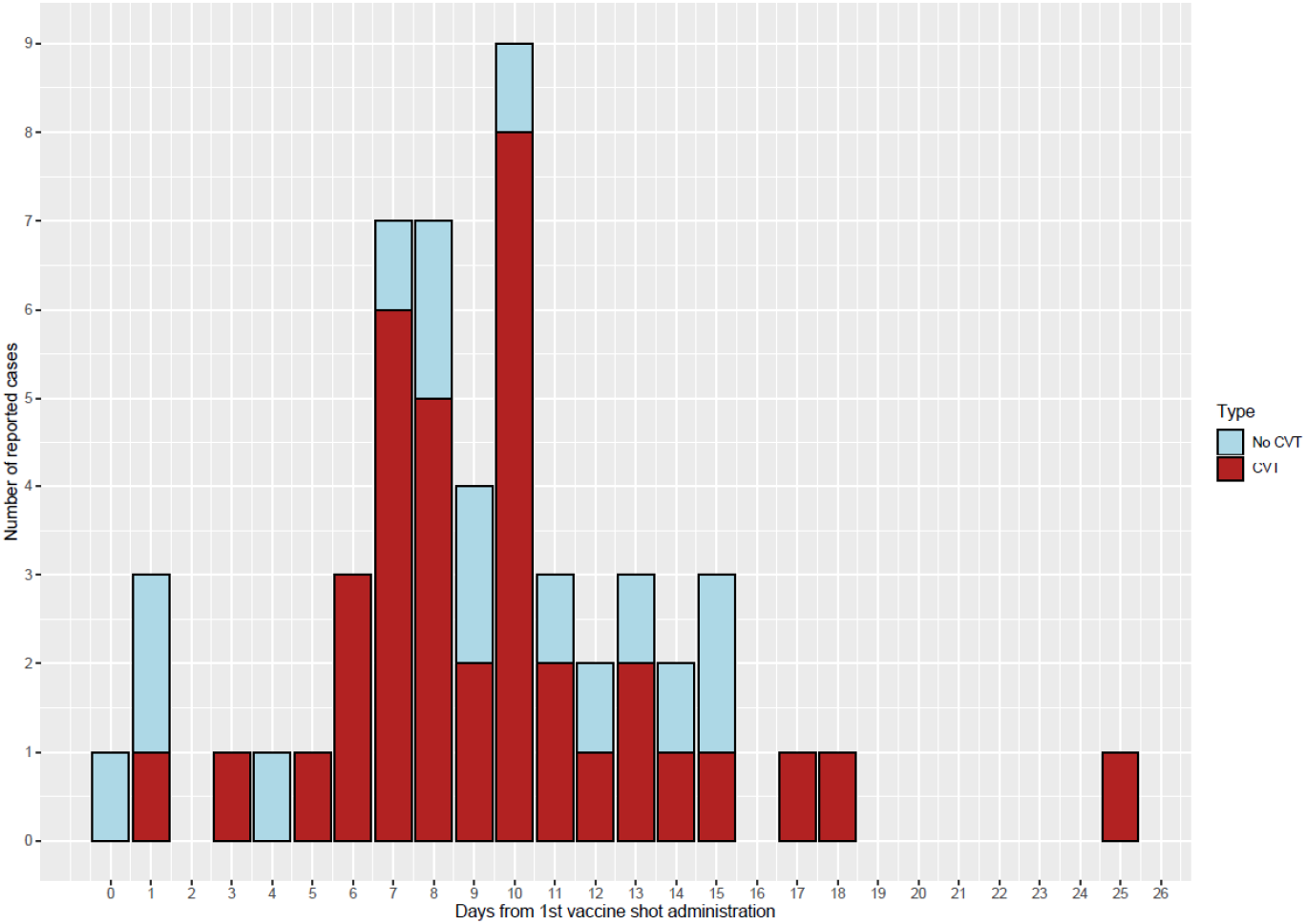
Days since first ChAdOx1 dose administration to neurological symptoms onset for CVT and non-CVT events.

With the prespecified VITT risk grading, we qualitatively investigated the adherence of the reported cerebrovascular events with the recently described syndrome of a vaccine-induced immunological syndrome leading to thrombocytopenia followed by thrombotic events. Overall, 4 (6.5%) had a risk score of 0, 24 (38.7%) had 1, 1 (1.6%) had 2, 5 (8.1%) had 2+, 4 (6.5%) had 3, and 24 (38.7%) had a score of 4. CVTs with a VITT risk score higher than 2 only occurred after vaccination with ChAdOx1. Among CVT cases, 20 (44.4%) scored 4 points, fulfilling all pre-defined criteria for the likelihood of vaccine association, and in 26 (57.8%) VITT was graded highly probable (Table 1).

In 3 patients, PF4 antibodies were positive, but the VITT function test was negative. All 28 patients with positive PF4 antibodies had received ChAdOx1 between 3 and 15 days before neurological symptoms. Among CVT cases, 22/31 (71.0%) had positive PF4 antibodies, 20/31 (64.5%) had positive VITT function test, 2/43 (4.7%) had relative thrombocytopenia, 26/43 (60.5%) had thrombocytopenia, and 23/33 (69.7%) had D-dimer levels above 500 µg/L (Table 1).

In addition to the CVT patients, nine cases with ischemic stroke were reported in this survey, eight (88.9%) of whom had received ChAdOx1 and one (11.1%) BNT162b2. Five (55.6%) of the 9 ischemic cases had a high (>2) VITT risk grade. Three (33.3%) fulfilled all 4 criteria of the VITT risk score (Table 1). In one of the two other cases with a score of three, the VITT function test was negative despite positive PF4 antibodies. In the second case with a score of three the thrombocytes were reduced to 152/nl but not below the threshold of 150/nl. Four patients with primary intracerebral bleeding without imaging signs of CVT were also reported, all after ChAdOx1 vaccination. One patient fulfilled all four pre-defined criteria of the VITT risk score (Table 1). In one other patient with primary intracerebral bleeding, PF4 antibodies and VITT function tests were not available, but severe thrombocytopenia and the typical time interval to the ChAdOx1 vaccination suggested a causal relationship.

Treatment was performed in 2/61 (3.3%) patients with plasmapheresis, 20/61 (32.8%) with intravenous high dose immunoglobulins and 4/61 (6.6%) with corticosteroids (Table 2). Anticoagulation was provided with heparin in 12/61 (19.7%), fraxiparin in 1/61 (1.6%), argatroban in 18/61 (29.5%), vitamin-K-antagonist 6/61 (9.8%), and direct oral anticoagulants in 9/61 (14.8%) cases. Eleven patients out of 60 (18.3%) died, of whom 9 had been vaccinated with ChAdOx1, and two had been vaccinated with BNT162b2. The distribution of the last available score (at discharge, death, or last available information if still hospitalized) on the modified Rankin scale by VITT risk score category is presented in Table 2.

**Table 2.** Treatments and modified Rankin scale of included cases with cerebral and central nervous system events within 31 days from Covid-19 vaccination by VITT risk group.

| Characteristics | VITT risk score $\leq 2$<br>(N=29) | VITT risk score $> 2$<br>(N=33) | Total (N=62) |
| --- | --- | --- | --- |
| **Plasmapheresis** |  |  |  |
| N-Miss | 0 | 1 | 1 |
| no | 29 (100.0%) | 30 (93.8%) | 59 (96.7%) |
| yes | 0 (0.0%) | 2 (6.2%) | 2 (3.3%) |
| **IVIG** |  |  |  |
| N-Miss | 0 | 1 | 1 |
| no | 28 (96.6%) | 13 (40.6%) | 41 (67.2%) |
| yes | 1 (3.4%) | 19 (59.4%) | 20 (32.8%) |
| **Cortisone** |  |  |  |
| N-Miss | 0 | 1 | 1 |
| no | 29 (100.0%) | 28 (87.5%) | 57 (93.4%) |
| yes | 0 (0.0%) | 4 (12.5%) | 4 (6.6%) |
| **Heparin sc or iv** |  |  |  |
| N-Miss | 0 | 1 | 1 |
| no | 23 (79.3%) | 26 (81.2%) | 49 (80.3%) |
| yes | 6 (20.7%) | 6 (18.8%) | 12 (19.7%) |
| **Heparin sc** |  |  |  |
| N-Miss | 0 | 1 | 1 |
| no | 23 (79.3%) | 28 (87.5%) | 51 (83.6%) |
| yes | 6 (20.7%) | 4 (12.5%) | 10 (16.4%) |
| **Heparin iv** |  |  |  |
| N-Miss | 0 | 1 | 1 |
| no | 29 (100.0%) | 28 (87.5%) | 57 (93.4%) |
| yes | 0 (0.0%) | 4 (12.5%) | 4 (6.6%) |
| **Fraxiparine** |  |  |  |
| N-Miss | 0 | 1 | 1 |
| no | 28 (96.6%) | 32 (100.0%) | 60 (98.4%) |
| yes | 1 (3.4%) | 0 (0.0%) | 1 (1.6%) |
| **Argatroban** |  |  |  |
| N-Miss | 0 | 1 | 1 |
| no | 26 (89.7%) | 17 (53.1%) | 43 (70.5%) |
| yes | 3 (10.3%) | 15 (46.9%) | 18 (29.5%) |
| **Vit_K** |  |  |  |
| N-Miss | 0 | 1 | 1 |
| no | 24 (82.8%) | 31 (96.9%) | 55 (90.2%) |
| yes | 5 (17.2%) | 1 (3.1%) | 6 (9.8%) |
| **DOAC** |  |  |  |
| N-Miss | 0 | 1 | 1 |
| no | 23 (79.3%) | 29 (90.6%) | 52 (85.2%) |
| yes | 6 (20.7%) | 3 (9.4%) | 9 (14.8%) |
| **last available mRS** |  |  |  |
| N-Miss | 1 | 1 | 2 |
| 0 | 10 (35.7%) | 7 (21.9%) | 17 (28.3%) |
| 1 | 13 (46.4%) | 3 (9.4%) | 16 (26.7%) |
| 2 | 1 (3.6%) | 2 (6.2%) | 3 (5.0%) |
| 3 | 0 (0.0%) | 1 (3.1%) | 1 (1.7%) |
| 4 | 1 (3.6%) | 3 (9.4%) | 4 (6.7%) |
| 5 | 1 (3.6%) | 7 (21.9%) | 8 (13.3%) |
| 6 | 2 (7.1%) | 9 (28.1%) | 11 (18.3%) |

VITT = vaccine-induced thrombocytopenic thrombosis
mRS = modified Ranking Scale (0 = no impairment to 6 death)

In total, we estimated an incidence rate of CVT within one month from first dose administration of 6.51 (95% CI, 4.43 to 9.25) per 100,000 person-years. This incidence rate was 17.91 (95% CI, 11.81 to 26.07) per 100,000 person-years for ChAdOx1, 1.32 (95% CI, 0.36 to 3.37) per 100,000 person-years for BNT162b2, and 0.00 (95% CI, 0.00 to 17.42) per 100,000 person-years for mRNA-1273. The incidence rate of CVT within one month from first ChAdOx1 dose administration was 23.53 (95% CI, 15.08 to 35.01) per 100,000 person-years for females and 6.16 (95% CI, 1.27 to 18.00) per 100,000 person-years for non-females. Incidence rates by age group, sex, and vaccine are reported in Table 3 and Figure 3.

**Table 3.** Incidence rates of CVT within one month (31 days) from first COVID-19 vaccine dose administration according to age group, sex, and vaccine type using data from nine States in Germany during the study period (January 1, 2021 to April 14, 2021).

| Age group (years) | Vaccine | Sex | Total doses administered | Estimated total first doses administered | Cases | Person-years | Rate per 100,000 person-years | 95% Confidence Interval |
| --- | --- | --- | --- | --- | --- | --- | --- | --- |
| <60 | ChAdOx1 | female | 1061837 | 1061837 | 20 | 82511.94 | 24.24 | 14.81 - 37.44 |
| <60 | ChAdOx1 | non-female | 449910 | 449910 | 3 | 33851.19 | 8.86 | 1.83 - 25.9 |
| <60 | BNT162b2 | female | 1354788 | 809594 | 2 | 55069.35 | 3.63 | 0.44 - 13.12 |
| <60 | BNT162b2 | non-female | 709366 | 451270 | 1 | 28359.32 | 3.53 | 0.09 - 19.65 |
| <60 | mRNA-1273 | female | 163110 | 105570 | 0 | 6568.7 | 0.00 | 0.00 - 56.16 |
| <60 | mRNA-1273 | non-female | 100068 | 70190 | 0 | 4024.96 | 0.00 | 0.00 - 91.65 |
| 60+ | ChAdOx1 | female | 423688 | 423688 | 4 | 19491.37 | 20.52 | 5.59 - 52.54 |
| 60+ | ChAdOx1 | non-female | 385100 | 385100 | 0 | 14858.32 | 0.00 | 0.00 - 24.83 |
| 60+ | BNT162b2 | female | 3171936 | 1889033 | 1 | 131407.46 | 0.76 | 0.02 - 4.24 |
| 60+ | BNT162b2 | non-female | 2154103 | 1304608 | 0 | 89202.54 | 0.00 | 0.00 - 4.14 |
| 60+ | mRNA-1273 | female | 161178 | 100096 | 0 | 6062.45 | 0.00 | 0.00 - 60.85 |
| 60+ | mRNA-1273 | non-female | 121116 | 75538 | 0 | 4515.66 | 0.00 | 0.00 - 81.69 |

**Figure 3.**
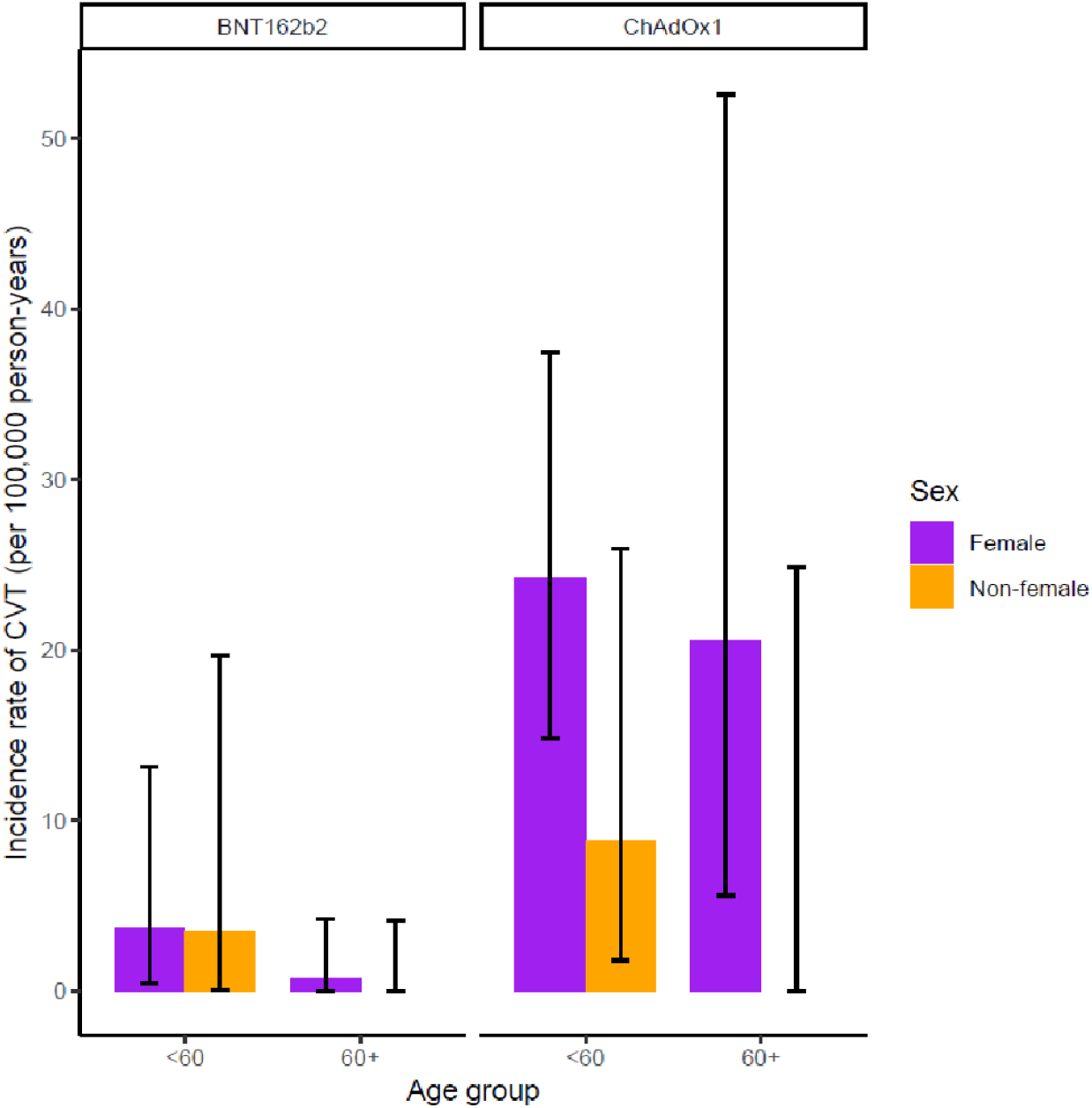
Incidence rate (95% confidence intervals) of CVT within one month (31 days) from first dose administration of vaccine against SARS-CoV-2 by vaccine type, sex, and age group.

In the model for CVT incidence within one month from first dose administration jointly considering age group, vaccine class, and sex, we estimated an adjusted incidence rate ratio of 9.68 (95% CI: 3.46 to 34.98, P <0.001) for ChAdOx1 compared to a mRNA-based vaccine, 3.14 (95% CI: 1.22 to 10.65, P=0.03) for females compared to non-females, and 2.14 (95% CI: 0.83 to 6.78, P=0.15) for those aged <60 compared with those aged 60 or more.

## Discussion

Our descriptive study from Germany identified 62 vascular cerebrovascular adverse events in close temporal relationship with a COVID-19 vaccination, of which 45 cases were CVT. We estimated an incidence rate of CVT within one month from first dose administration of 17.9 per 100,000 person-years for the ChAdOx1 vaccine and 1.3 per 100,000 person-years for BNT162b2. Before the COVID-19 pandemic, the incidence rate of CVT has been estimated between 0.22 – 1.75 per 100,000 person-years in four European countries, Australia, Iran, and Hong Kong^7–9^. This corresponds to an over 10-fold higher CVT incidence rate in patients who received a first ChAdOx1 vaccine shot compared with the highest estimate of CVT incidence rate from empirical data. The incidence rate of a CVT event after first dose COVID-19 vaccination was statistically significantly higher for ChAdOx1 (rate ratio: 9.68, 3.46 to 34.98) compared to mRNA-based vaccines and for females (3.14, 1.22 to 10.65) compared to non-females. In our data, the association between age group (60+, <60) and CVT incidence was not statistically significant after accounting for sex and vaccine class.

Comparisons with other countries and settings are challenging, as the probability of receiving a specific vaccination differs by age, sex, profession, and other factors. The number of people diagnosed in the UK with CVT after receiving the ChAdOx1 vaccine was reported by the UK Medicines & Healthcare products Regulatory Agency (MHR) as 22 cases among 18 million people have received the vaccine^17^ However, these numbers were updated by the MHR by April 28, 2021. Now, 242 cases of major thromboembolic events with concurrent thrombocytopenia following vaccination with ChAdOx1 were reported, including 93 cases of CVT^18^. Up to the same time point, 22.6 million Britains had received their first dose and 5.9 million their second dose of ChAdOx1. Of the 242 cases, 161 occurred in the age group below and 67 in the age group above 60. In 14 cases, the age remained unknown.

A population-based cohort study using data from 281,264 individuals vaccinated with ChAdOx1 in Denmark and Norway reported a standardized morbidity ratio for cerebral venous thrombosis of 20.25, corresponding to 7 observed events versus 0.3 expected ones and an excess of 2.5 events per 100,000 vaccinations^19^. While there was no overall increase in the arterial event group, the rate of intracerebral hemorrhage was increased, with a standardized morbidity ratio of 2.33^19^.

CVT is a very rare disease, and it is unlikely that the higher incidence rate among vaccinated is purely the product of chance. The identification of antibodies against thrombocytes in a high percentage of our patients in whom the test results were available is another strong argument for a causal relationship. Understandably, the recommended treatment of CVT beyond anticoagulation is the use of IVIG or plasmapheresis.

While our survey focused on collecting information about CVT cases in temporal relationship with COVID-19 vaccination, our questionnaire also allowed the reporting of other neurological diagnoses. Interestingly, 5 cases with embolic ischemic stroke and a VITT score of >2 without signs of CVT were reported. In four of them, thrombotic occlusion of the middle cerebral artery, the internal carotid artery and/or recurrent thrombotic material in duplex ultrasound were reported. This is similar to Heparin-induced thrombopenia, in which arterial thrombosis occurs, as well, and at a ratio of 1:4.3 compared with venous thrombosis^20^. In addition, two primary intracranial hemorrhages with a VITT risk score >2 without a detectable sign of CVT were reported.

The conclusions with respect to our hypotheses are as follows:

i. Individuals in Germany were vaccinated with ChAdOx1, mRNA-1273, and BNT162b2. In our study, CVTs with a VITT risk score >2 only occurred after vaccination with ChAdOx1. Our results suggest that VITT-induced antibodies against PF4 do not cross-react with the spike protein of SARS-CoV-2^21^. A recent report of an individual who developed a CVT associated with severe thrombopenia at 14 days after immunization with Ad26.COV2.S suggests that VITT-associated thrombotic events may be associated with adenovirus vector-based vaccines directed against the SARS-CoV-2 spike protein^22^. Since ChAdOx1 is based on a chimpanzee and Ad26.COV2.S on a human adenovirus vector, and they differ in their spike protein inserts, there was hope that a VITT is restricted to ChAdOx1.
ii. We confirm that CVT within one month from first dose administration occur at a higher rate in females compared to non-females, accounting for age group and vaccine class. Indeed, the adjusted rate ratio was equal to 3.1 (95% CI: 1.2 – 10.6). However, the rate of CVT occurring within one month from first dose administration once accounted for sex, and vaccine class did not significantly differ (p-value=0.15) between individuals younger than 60 years and individuals that are 60 years or older.
iii. We confirm that most of the patients with a CVT at one to 16 days after vaccination with ChAdOx1, who have thrombocytopenia, also have VITT.
iv. VITT-mediated cerebral vascular events (VITT risk score >2) were not restricted to CVT but were also observed in cases of primary cerebral ischemia (n=5) and intracerebral hemorrhage (n=2).

Currently, several questions remain unanswered. First, it is unclear how many patients develop antibodies against PF4 after vaccination with ChAdOx1 (and potentially Ad26.COV2.S) without thrombotic complications. Therefore, the risk of re-exposure to the vaccine in conjunction with the second vaccination cannot be estimated. Second, although the platelet activation of VITT is heparin-independent, it is unknown whether heparin therapy aggravates VITT in analogy to the clinical syndrome of autoimmune heparin-induced thrombocytopenia. Hence, non-heparin anticoagulants are recommended for the treatment of VITT-related CVT. Although venous and less common arterial thromboses have also been reported outside the central nervous system in VITT^12,21^, it remains unclear why vessels of the central nervous system are primarily affected.

Our data cannot serve and should not be interpreted as a recommendation for the vaccination strategy to be implemented. While we believe this article provides important information to inform such a decision, we only quantified the incidence of cerebrovascular events following vaccination by sex, age group, and vaccine type in nine German states. The decision on which vaccination strategy is best in a specific context depends not only on the risks of the vaccination but also on its benefits, with respect to possible and available alternative strategies. Specifically, it needs to be emphasized that VITT is a very rare event and that the risk-benefit ratio of vaccination against SARS-CoV-2 needs to be considered. Other factors to be taken into account for estimating an overall risk-benefit ratio include the risk of cerebral blood clots from COVID-19 disease^23,24^, the existence, and availability of alternative vaccines.

Strengths of our study include the standardized collection of patient data with cerebral outcomes within a reasonable time period after COVID-19 vaccination from almost all Departments of Neurology of German university hospitals (which represent the tertiary care centers in Germany). As of March 31, 2021, a total of 31 cases of CVT after ChAdOx1 vaccination in the whole country have been reported^25^. In our study, we reported 37 confirmed CVT cases after ChAdOx1 vaccination as of April 15, suggesting a high level of coverage in the case ascertainment. Another strength of the study is that each case was evaluated by four neurologists and one coagulation specialist who discussed all aspects of the provided clinical information. To approximate the incidences, we used official data of vaccinated people in Germany and had information on the age, sex and vaccine type distribution from 9 of the 16 States in Germany.

Limitations of our study include that we only collected information from neurological departments, and patients may have been treated at other departments or died without reaching a hospital. We could not collect the brain imaging data to validate the diagnosis of CVT and other cerebral events. Moreover, our main disease of interest was CVT after COVID-19 vaccination. While we can be confident, also thanks to public discussions about CVT as a consequence of the ChAdOx1 vaccine, that we have high coverage for this disease, this does not hold true for the other cerebral diseases. Almost certainly, the cases of ischemic stroke or cerebral hemorrhage reported in this survey represented a very selected subset of all cases, probably reported because of a strong suspicion of a link with the COVID-19 vaccination. We did not have data on the age, sex, and vaccine type distribution of vaccinated people from all of Germany but only from 9 of its 16 states. In the Robert Koch-Institute dataset, the vaccine shots administered by general practitioners are not included, probably leading to an overestimation of the true risk.

Furthermore, we had to compute the overall number of person-years spent at risk by subgroup, relying on a crude approximation of the number of first doses administered. Lastly, we cannot exclude that in this retrospective survey some transferred data were misclassified or differentially missing. Two of the cases included in this report were also presented in the first description of VITT by Greinacher and colleagues^13^.

### Implications

Findings of our study imply further careful considerations in the administration of ChAdOx1, especially for women, and risk-benefit considerations when considering this vector-based vaccination by age. In addition, continued registration of all cerebrovascular events after vaccination and all rare cerebral venous thromboses in a standardized and validated manner is important for properly evaluating the risk of these events after COVID-19 vaccination.

## Data Availability

Due to the votes that we received for ethics and data protection and privacy conformity for this survey, we will not be able to share data that contain information about the center, the age and the sex of the patient.

## Acknowledgments

We thank the Robert Koch-Institute for providing data on administered vaccine shots in Germany.

## Authors contributions

The corresponding author attests that all listed authors meet authorship criteria and that no others meeting the criteria have been omitted.

JBS: conception and design, clinical neurological expertise, supervision, drafting of the manuscript, and critical comments and approved the final version of the manuscript.

PB: conception and design, clinical neurological expertise, critical comments, and approved the final version of the manuscript.

HCD: clinical neurological and epidemiological expertise, drafting of the manuscript, critical comments, and approved the final version of the manuscript.

CG: conception and design, clinical neurological expertise, supervision, critical comments, and approved the final version of the manuscript.

AG: hematological expertise and analysis, supervision, critical comments, and approved the final version of the manuscript.

CK: clinical neurological expertise, critical comments, and approved the final version of the manuscript.

GCP: clinical neurological expertise, critical comments, and approved the final version of the manuscript.

MP: epidemiological and statistical expertise, data analysis, creation of the figures, drafting of the manuscript, critical comments, and approved the final version of the manuscript.

SP: clinical neurological expertise, critical comments, and approved the final version of the manuscript.

RR: conception and design, technical support for data ascertainment, data management, medical informatics expertise, critical comments, and approved the final version of the manuscript.

HS: clinical neurological expertise, critical comments, and approved the final version of the manuscript.

TT: hematological expertise and analysis, critical comments, and approved the final version of the manuscript.

TK: conception and design, epidemiological expertise, supervision of the analyses, drafting of the manuscript, critical comments and approved the final version of the manuscript.

All authors had access to the data. JBS is guarantor and accepts full responsibility for the work and the conduct of the study.

## PB serves as editor-in-chief for DGNeurologie

HCD reports outside of the present study: received honoraria for participation in clinical trials, contribution to advisory boards or oral presentations from: Abbott, BMS, Boehringer Ingelheim, Daiichi-Sankyo, Novo-Nordisk, Pfizer, Portola and WebMD Global. Financial support for research projects was provided by Boehringer Ingelheim. HCD received research grants from the German Research Council (DFG), German Ministry of Education and Research (BMBF), European Union, NIH, Bertelsmann Foundation, and Heinz-Nixdorf Foundation. HCD serves as editor of Neurologie up2date, Info Neurologie & Psychiatrie, Arzneimitteltherapie, as co-editor of Cephalalgia and on the editorial board of Lancet Neurology. HCD chairs the Treatment Guidelines Committee of the German Society of Neurology and contributed to the EHRA and ESC guidelines for the treatment of AF.

CG received (outside the submitted work) funding from German Research Council (DFG), European Union, Federal Ministry of Education and Research (BMBF), German Statutory Pension Insurance Scheme (RV Nord), National Innovation Fund, Wegener Foundation, and Schilling Foundation. CG reports (outside the submitted work) personal fees from Amgen, Boehringer Ingelheim, Daiichi Sankyo, Abbott, Prediction Biosciences, Novartis, and Bayer.

AG reports grants from Deutsche Forschungsgemeinschaft, personal fees and non-financial support from Aspen, grants from Ergomed, grants and non-financial support from Boehringer Ingelheim, personal fees from Bayer Vital, grants from Rovi, grants from Sagent, personal fees from Chromatec, personal fees and nonfinancial support from Instrumentation Laboratory, grants and personal fees from Macopharma, grants from Portola, grants from Biokit, personal fees from Sanofi-Aventis, grants from Fa. Blau Farmaceutics, grants from Prosensa/Biomarin, grants and other from DRK-BSD NSTOB, grants from DRK-BSD Baden-Wuertemberg/Hessen, personal fees and non-financial support from Roche, personal fees from GTH e.V., grants from Deutsche Forschungsgemeinschaft, outside the submitted work. In addition, AG reports having a patent, Application no. 2021032220550000DE, pending.

CK serves as a medical advisor on genetic testing reports to Centogene and on the scientific advisory board of Retromer Therapeutics and as an Associate Editor for Annals of Neurology.

MP reports being partially funded by a research grant from Novartis Pharma for a self-initiated research project, unrelated to this work, on migraine remission. MP further reports being awarded a research grant from the Center for Stroke Research Berlin (private donations) for a self-initiated project unrelated to this work on causal diagrams.

SP received (all outside the submitted work) research grants from BMS/Pfizer, Daiichi Sankyo, European Union, German Federal Joint Committee Innovation Fund, and German Federal Ministry of Education and Research, and speakers honoraria/consulting fees from AstraZeneca, Bayer, Boehringer-Ingelheim, BMS/Pfizer, Daiichi Sankyo, Portola, and Werfen.

RR reports outside of the submitted work to have received research funds from the German Ministry of Education and Research, German Joint Committee (GBA) and the German Ministry of Health.

HS reports outside of the submitted work to have received speaker’s honoraria from Bayer, Boehringer Ingelheim, and Teva.

TT reports grants from Deutsche Forschungsgemeinschaft, during the conduct of the study; personal fees and other from Bristol Myers Squibb, personal fees and other from Pfizer, personal fees from Bayer, personal fees and other from Chugai Pharma, other from Novo Nordisk, personal fees from Novartis, other from Daichii Sankyo, outside the submitted work.

TK reports outside of the submitted work to have received research funds from the German Joint Committee and the German Ministry of Health; he has received personal compensation from Eli Lilly & Company, Teva, Total S.E., and the BMJ.

## Data sharing

Due to the votes that we received for ethics and data protection, and privacy conformity for this survey, we will not be able to share data that contain information about the center, the age, and the sex of the patient.

## Dissemination

Upon request, the data will be available for policymakers and government bodies.

The guarantor (JBS) affirms that the manuscript is an honest, accurate, and transparent account of the study being reported; that no important aspects of the study have been omitted; and that any discrepancies from the study as originally planned (and, if relevant, registered) have been explained.

## DGN Covid-19 Vaccination Study Group

Prof. Dr. Angelika Alonso, Medizinische Fakultät Mannheim, Universität Heidelberg;

Prof. Dr. Thorsten Bartsch, Klinik für Neurologie, Universitätsklinikum Schleswig-Holstein Kiel

Dr. Christoph Baumsteiger, Neurologie und Neurophysiologie, LVR Klinik Bedburg-Hau

Dr. med. Felix Bode, Klinik für Neurologie, Uniklinik Bonn

Dr. Hakan Cangür, Neurologische Klinik, Klinikum Wolfsburg

Prof. Michael Daffertshofer, Neurologie, Klinikum Mittelbaden, Rastatt

PD Dr. med. Manuel Dafotakis, Neurologische Klinik, Uniklinik, RWTH Aachen

Prof. Dr. med. Marianne Dieterich, LMU München, Klinik für Neurologie

PD Dr. med. Ralf Dittrich Niels, Stensen Kliniken, Marienhospital Osnabrück, Klinik für Neurologie

Dr. Friederike Fabian, Neurologie, Marienhospital, Stuttgart

Dr. med. Mathias Fousse, Uniklinik des Saarlandes, Abteilung für Neurologie

Dr. Jana Godau, Klinikum Kassel, Neurologische Klinik

Prof. Dr. Martin Grond, Kreisklinikum Siegen, Klinik für Neurologie

Dr. med. Albrecht Günther, Hans-Berger-Klinik für Neurologie, Universitätsklinikum Jena, Am Klinikum 1, 07747 Jena

Prof. Dr. med. Alexander Gutschalk, Neurologische Klinik, Universitätsklinikum Heidelberg

Prof. Dr. med. Georg Hagemann, Helios Klinikum Berlin-Buch, Klinik für Neurologie;

Dr. med. Corinna Hartmann, Helios Klinik Lengerich

Prof. Rüdiger Hilker-Roggendorf, Knappschaftskrankenhaus Recklinghausen, Klinik für Neurologie

Prof. Dr. med. Günter Höglinger, Klinik für Neurologie, Medizinische Hochschule Hannover

Dr. Benno Ikenberg; Neurologische Klinik, Klinikum rechts der Isar der TU München)

Dr. med. Fatme Seval Ismail, Department of Neurology, University Hospital Knappschaftskrankenhaus Bochum, Ruhr University Bochum, Germany

PD Dr. med. Sarah Jesse, Ulm University, Department of Neurology, Oberer Eselsberg 45, 89081 Ulm

PD Dr. Bernd Kallmünzer, Neurologische Universitätsklinik Erlangen.

Prof. Dr. med. Rolf Kern, Klinik für Neurologie, Klinikverbund Allgäu

Dr. med. Martin Klietz, Klinik für Neurologie, Medizinische Hochschule Hannover

Dr. med. Samuel Knauß, Klinik für Neurologie, Charité, Berlin

PD Dr. med. Benjamin Knier, Neurologische Klinik, Klinikum rechts der Isar, Technische Universität München, München, Deutschland

Prof. Dr. Volker Limmroth, Neurologische Klinik, Kliniken der Stadt Köln

Dr. med. Annerose Mengel, Department of Neurology and Stroke, Universitätsklinikum Tübingen

Dr. Johannes Meyne, UKSH, Campus Kiel, Klinik für Neurologie Dr. med. Martin Morgenthaler, Westpfalz-Klinikum GmbH

Dr. med. Matthias Müller, Abteilung für Neurologie, AK Nord Heidberg

Prof. Dr. med. Simon Nagel, Neurologische Klinik, Universitätsklinikum Heidelberg

PD Dr. Oezguer A. Onur, University of Cologne, Faculty of Medicine and University Hospital Cologne, Department of Neurology, 50937 Cologne, Germany

Dr. med. Johann Pelz, Klinik und Poliklinik für Neurologie, Universitätsklinikum Leipzig, Leipzig

Dr. Johannes Plenge, USKH, Klinik für Neurologie

PD Dr. Sven Poli, Universitätsklinikum Tübingen

Dr. med. Christian Roth, Neurologie, DRK-Kliniken Nordhessen, Kassel

Prof. Dr. med. Joachim Röther, Asklepios Klinik Altona

Dr. med. Christian Saß, Asklepios Klinikum Harburg, Neurologie

PD Dr. med. Silvia Schönenberger, Neurologische Klinik, Universitätsklinikum Heidelberg

Dr. med. Roger Schubert, Klinik für Neurologie, SRH-Waldklinikum Gera

Dr. med. Ole Simon, Universitätsklinikum Gießen und Marburg, Standort Marburg

Ina Specht, Agaplesion Diakonieklinikum Rotenburg, Neurologische Klinik

Dr. Anne Sperfeld, Neurologische Klinik Altscherbitz, Schkeuditz

PD Dr. Annette Spreer, Klinikum Braunschweig

Prof. Dr. med. Andreas Steinbrecher; Klinik für Neurologie, Helios Klinikum Erfurt

Dr. med. Jochen Steiner, Universitätsklinikum Tübingen;

Dr. Henning Stetefeld, Uniklinik Köln der Universität zu Köln

Prof. Dr. George Trendelenburg, Klinikum Itzehoe & Universitätsmedizin Göttingen

Dr. med. Nils Bijan Vatankhah, Diako Flensburg;

Dr. med. Christoph Michael Wahl; Neurologie, Klinikum Kempten

Dr. med. Katja Wartenberg, Klinik und Poliklinik für Neurologie, Universitätsklinikum Leipzig, Leipzig

Prof. Dr. med. Karsten Witt, Dept. of Neurology and Center of Neurosensory Sciences, University Oldenburg, Germany

PD Dr. med. Matthias Wittstock, Klinik und Poliklinik für Neurologie, Universitätsmedizin Rostock

Dr. med. Björn Wolf

PD Dr. Joachim Wolf, Diakonissenkrankenhaus Mannheim

Dr. med. Julian, Zimmermann, Uniklinik Bonn Klinik für Neurologie

## References

1. Ramasamy MN, Minassian AM, Ewer KJ, et al. Safety and immunogenicity of ChAdOx1 nCoV-19 vaccine administered in a prime-boost regimen in young and old adults (COV002): a single-blind, randomised, controlled, phase 2/3 trial. Lancet 2021;396(10267):1979–1993.

2. Voysey M, Clemens SAC, Madhi SA, et al. Safety and efficacy of the ChAdOx1 nCoV-19 vaccine (AZD1222) against SARS-CoV-2: an interim analysis of four randomised controlled trials in Brazil, South Africa, and the UK. Lancet 2021;397(10269):99–111.

3. Polack FP, Thomas SJ, Kitchin N, et al. Safety and Efficacy of the BNT162b2 mRNA Covid-19 Vaccine. New Engl J Med 2020;

4. Baden LR, Sahly HME, Essink B, et al. Efficacy and Safety of the mRNA-1273 SARS-CoV-2 Vaccine. New Engl J Med 2020;384(5):403–416.

5. Sadoff J, Gray G, Vandebosch A, et al. Safety and Efficacy of Single-Dose Ad26.COV2.S Vaccine against Covid-19. New Engl J Med 2021;

6. Robert-Koch-Institut. Impfquotenmonitoring [Internet]. 2021;[cited 2021 Apr 18] Available from: https://www.rki.de/DE/Content/InfAZ/N/Neuartiges_Coronavirus/Daten/Impfquotenmonitoring.xlsx?__blob=publicationFileThese

7. Ruuskanen JO, Kytö V, Posti JP, et al. Cerebral Venous Thrombosis: Finnish Nationwide Trends. Stroke 2021;52(1):335–338.

8. Kristoffersen ES, Harper CE, Vetvik KG, et al. Incidence and Mortality of Cerebral Venous Thrombosis in a Norwegian Population. Stroke 2020;51(10):3023–3029.

9. Ferro JM, Correia M, Pontes C, et al. Cerebral Vein and Dural Sinus Thrombosis in Portugal: 1980–1998. Cerebrovasc Dis 2001;11(3):177–182.

10. Idiculla PS, Gurala D, Palanisamy M, et al. Cerebral Venous Thrombosis: A Comprehensive Review. Eur Neurol 2020;83(4):369–379.

11. Marjot T, Yadav S, Hasan N, et al. Genes Associated With Adult Cerebral Venous Thrombosis. Stroke 2011;42(4):913–918.

12. Schultz NH, Sørvoll IH, Michelsen AE, et al. Thrombosis and Thrombocytopenia after ChAdOx1 nCoV-19 Vaccination. New Engl J Med 2021;

13. Greinacher A, Thiele T, Warkentin TE, et al. Thrombotic Thrombocytopenia after ChAdOx1 nCov-19 Vaccination. New Engl J Med 2021;

14. Scully M, Singh D, Lown R, et al. Pathologic Antibodies to Platelet Factor 4 after ChAdOx1 nCoV-19 Vaccination. New Engl J Med 2021;

15. Wise J. Covid-19: European countries suspend use of Oxford-AstraZeneca vaccine after reports of blood clots. BMJ 2021;372:n699.

16. Østergaard SD, Schmidt M, Horváth-Puhó E, et al. Thromboembolism and the Oxford–AstraZeneca COVID-19 vaccine: side-effect or coincidence? Lancet 2021;

17. MHR. Coronavirus vaccine - weekly summary of Yellow Card reporting [Internet]. n.d.;[cited 2021 March 30] Available from: https://www.gov.uk/government/publications/coronavirus-covid-19-vaccine-adverse-reactions/coronavirus-vaccine-summary-of-yellow-card-reporting

18. MHR. Coronavirus vaccine - weekly summary of Yellow Card reporting Updated May 6 2021 [Internet]. n.d.;Available from: https://www.gov.uk/government/publications/coronavirus-covid-19-vaccine-adverse-reactions/coronavirus-vaccine-summary-of-yellow-card-reporting

19. Pottegård A, Lund LC, Karlstad Ø, et al. Arterial events, venous thromboembolism, thrombocytopenia, and bleeding after vaccination with Oxford-AstraZeneca ChAdOx1-S in Denmark and Norway: population based cohort study. BMJ 2021;373:n1114.

20. Warkentin TE, Kelton JG. JG 14-year study of heparin-induced thrombocytopenia. Am J Medicine 1996;101(5):502–507.

21. Greinacher A, Thiele T, Warkentin TE, et al. A Prothrombotic Thrombocytopenic Disorder Resembling Heparin-Induced Thrombocytopenia Following Coronavirus-19 Vaccination. 2021;

22. Muir K-L, Kallam A, Koepsell SA, Gundabolu K. Thrombotic Thrombocytopenia after Ad26.COV2.S Vaccination. New Engl J Med 2021;

23. Taquet M, Husain M, Geddes JR, et al. Cerebral venous thrombosis: a retrospective cohort study of 513,284 confirmed COVID-19 cases and a comparison with 489,871 people receiving a COVID-19 mRNA vaccine [Internet]. 2021;Available from: https://osf.io/a9jdq/

24. Torjesen I. I-19: Risk of cerebral blood clots from disease is 10 times thatfrom vaccination, study finds [Internet]. BMJ 2021;373:n1005 n.d.;Available from: http://dx.doi.org/10.1136/bmj.n1005

25. Dyer O. O-19: EMA defends AstraZeneca vaccine as Germany and Canada halt rollouts. BMJ. 2021 Apr 1;373:n883.

